# COVID-19 dynamics in an Ohio prison

**DOI:** 10.1101/2021.01.14.21249782

**Authors:** Wasiur R. KhudaBukhsh, Sat Kartar Khalsa, Eben Kenah, Grzegorz A. Rempala, Joseph H. Tien

## Abstract

**Importance:** Incarcerated individuals are a vulnerable population for severe acute respiratory syndrome coronavirus 2 (SARS-CoV-2) infection. Understanding SARS-CoV-2 dynamics in prisons is crucial for curbing transmission both within correctional facilities and in the surrounding community.

**Objective:** The purpose of this study was to identify transmission scenarios that could underly rapid, widespread SARS-CoV-2 infection among inmates in Marion Correctional Institution (MCI).

**Design:** Publicly available data reported by the Ohio Department of Rehabilitation and Corrections (ODRC) was analyzed using mathematical and statistical models.

**Setting:** We consider SARS-CoV-2 transmission dynamics among MCI inmates prior to and including April 16, 2020.

**Participants:** This study uses de-identified, publicly available SARS-CoV-2 test result data for MCI inmates.

**Exposures:** Inmates at MCI were considered exposed to potential infection with SARS-CoV-2.

**Main outcome and measures:** Results from mass testing conducted on April 16, 2020 were analyzed together with time of first reported SARS-CoV-2 infection among MCI inmates.

**Results:** Rapid, widespread infection of MCI inmates was reported, with nearly 80% of inmates infected within three weeks of first reported inmate case. These data are consistent with i) a basic reproduction number greater than 14, together with a single initially infected inmate, ii) an initial super-spreading event resulting in several hundred initially infected inmates, together with a basic reproduction number of approximately three, and iii) earlier undetected circulation of virus among inmates prior to April.

**Conclusions and relevance:** Mass testing data are consistent with extreme transmissibility, super-spreading events, or undetected circulation of virus among inmates. All scenarios consistent with these data attest to the vulnerabilities of prisoners to COVID-19.

**Key points:** *Question:* To identify transmission characteristics consistent with timing and extent of SARS-CoV-2 infection among inmates in Marion Correctional Institution.

*Findings:* Mathematical and statistical modeling finds three scenarios that are consistent with the observed widespread infection in Marion Correctional Institution: i) very high transmissibility corresponding to a basic reproduction number in the double digits, ii) an initial super-spreading event involving exposure of several hundred inmates, iii) undetected circulation of virus prior to the first documented case among inmates.

*Meaning:* High transmissibility, super-spreading events, and challenges with disease surveillance all attest to the vulnerabilities of prison populations to SARS-CoV-2.

## 1 Introduction

Incarcerated individuals are a vulnerable population for severe acute respiratory syndrome coronavirus 2 (SARS-CoV-2). Factors contributing to SARS-CoV-2 transmission in prisons include shared housing, crowding, inability to social distance, challenges for hygiene, and more. Transmission can be extensive: as of November 6, 2020, various testing protocols across states and jurisdictions have identified 169,286 COVID-19 cases among prisoners and 38,438 cases among prison staff in the United States [1]. Outbreak sizes within facilities can be high: infections in more than 80% of prisoners at Marion Correctional Institution (MCI) in Ohio have been identified^1^, and similarly high levels of infection have been observed in correctional facilities under other jurisdictions [3]. The vulnerability of prisoners to COVID-19 and the potential for the penal system to serve as hot spots for transmission have been noted by many authors [4–8].

These outbreak sizes indicate widespread SARS-CoV-2 transmission. Perhaps even more remarkable is the apparent *intensity* of transmission within prisons. Here we analyze publicly available COVID-19 time series data from the Ohio Department of Rehabilitation and Corrections (ODRC). This analysis highlights the extreme potential for transmission in prisons, as well as the need for high quality, publicly available data on COVID-19 in correctional facilities.

## 2 Data and Methods

### Case data

Mass RT-PCR testing at MCI of all inmates and partial testing of staff was conducted on April 16, 2020. Total number of inmates and number of inmates and staff at MCI testing positive for SARS-CoV-2 over time were obtained from public ODRC reports [2]. To account for lags in data entry, we accumulate the cases reported at MCI over April 16-23, 2020 as a single mass testing data point.

### Mathematical model

We use a susceptible-exposed-infectious-recovered (SEIR) framework to model SARS-CoV-2 dynamics in MCI. Under the standard SEIR model, the proportions of individuals in the susceptible (*S*_*t*_), exposed (*E*_*t*_), infectious (*I*_*t*_), and recovered (*R*_*t*_) compartments (at time *t*) satisfy the following system of differential equations:

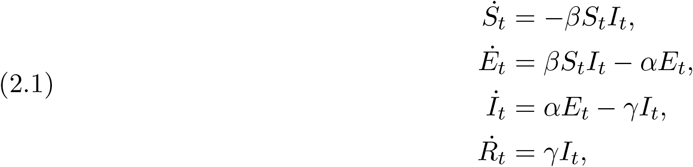

where the positive parameters *β, α*, and *γ* denote the infection rate, the incubation rate, and the recovery rate.

### Statistical analysis

We derive a likelihood function for observing *n* positives out of *N* incarcerated individuals on day *u* as follows. Using the Dynamic Survival Analysis approach of [9], we treat *S*_*t*_ as an improper survival function. Given observation up to time *T >* 0, the time *T*_*E*_ that an individual becomes infected and enters the *E* compartment follows the conditional probability density function

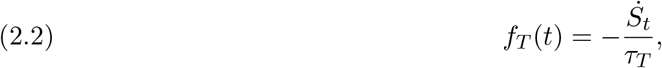

where *τ*_*T*_ = 1 − *S*_*T*_. The time *T*_*I*_ to becoming infectious has the conditional density

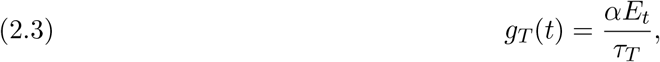

and the recovery time *T*_*R*_ has the conditional density

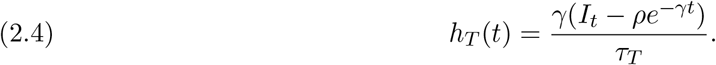

Note that the random variables *T*_*E*_, *T*_*I*_ − *T*_*E*_, and *T*_*R*_ − *T*_*E*_ are mutually independent, and that *T*_*I*_ − *T*_*E*_ and *T*_*R*_ − *T*_*E*_ are exponentially distributed with rates *α* and *γ*.

Mass testing yields the number of individuals who test positive and the total number of tests administered on the day of mass testing. To use these data, let *T*_*N*_ denote the time when virus first becomes undetectable in an individual. We then describe the epidemic process by the pair of random variables (*T*_*E*_, *T*_*N*_). Let *ε* = *T*_*N*_ − *T*_*E*_. Then, the conditional probability of an individual testing positive on the day of the mass testing (at time *u*) is given by

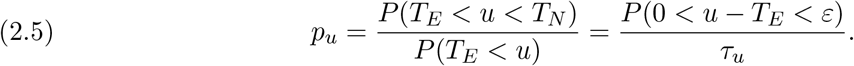

We fix *ε* = 21 days, corresponding to detectable virus for three weeks following an individual becoming infectious [10, 11]. We set 1*/α* = 5.1*/* log(2) days (corresponding to a median incubation period of 5.1 days [12]), and assume a mean infectious period 1*/γ* of 5.6 days [13].

If *n* out of *N* individuals test positive on the day of the mass testing *u*, the likelihood function is given by

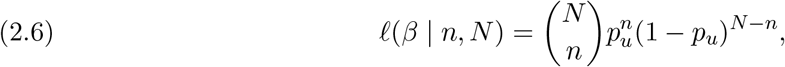

with the probability of testing positive *p*_*u*_ as described in (2.5).

## 3 Results

### Reported outbreak time course

According to ODRC reports [2], the first identified COVID-19 case at MCI was an infected staff member on March 29. Following this initial primary case, precautions such as cohorting and modified movement were enacted in order to restrict mixing and reduce transmission.^2^ The first COVID-19 case among inmates was identified on April 3. Mass RT-PCR testing of all inmates and partial testing of staff was conducted on April 16. By April 20, SARS-CoV-2 infection had been identified in 79% (1950/2453) of inmates and 35% (154/446) of staff.^3^ A time series of COVID-19 cases at MCI is shown in Figure 1.

**Figure 1:**
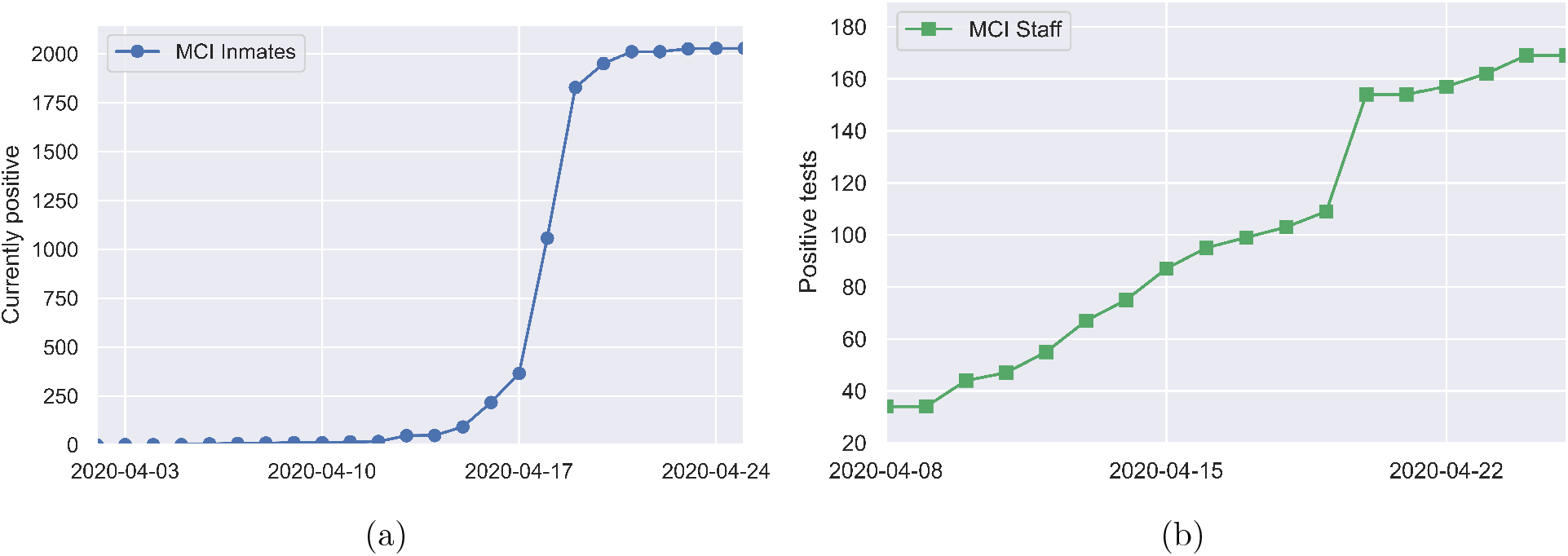
Positive tests over time among (a) inmates and (b) staff at MCI, as reported in [2].

### Basic reproduction number and initial exposure size

To examine what values for the basic reproduction number *ℛ*_0_ are consistent with such rapid spread, we turn to the SEIR model (system (2.1)). In order for mass screening to identify 80% of the population as positive for COVID-19 on April 16, at least 80% of the population must have been infected by that date. Figure 2 shows the time needed to infect 80% of the population in the SEIR model as a function of the basic reproduction number (*ℛ*_0_) and the initial number of exposed individuals (*E*_0_). While *ℛ*_0_ values of two or larger are able to eventually infect 80% or more of the population, for modest *ℛ*_0_ values this can take on the order of months. Reproduction numbers larger than 14 are needed before outbreaks originating from a single exposed individual are able to generate infection levels in a three week span that are consistent with those reported for MCI.

**Figure 2:**
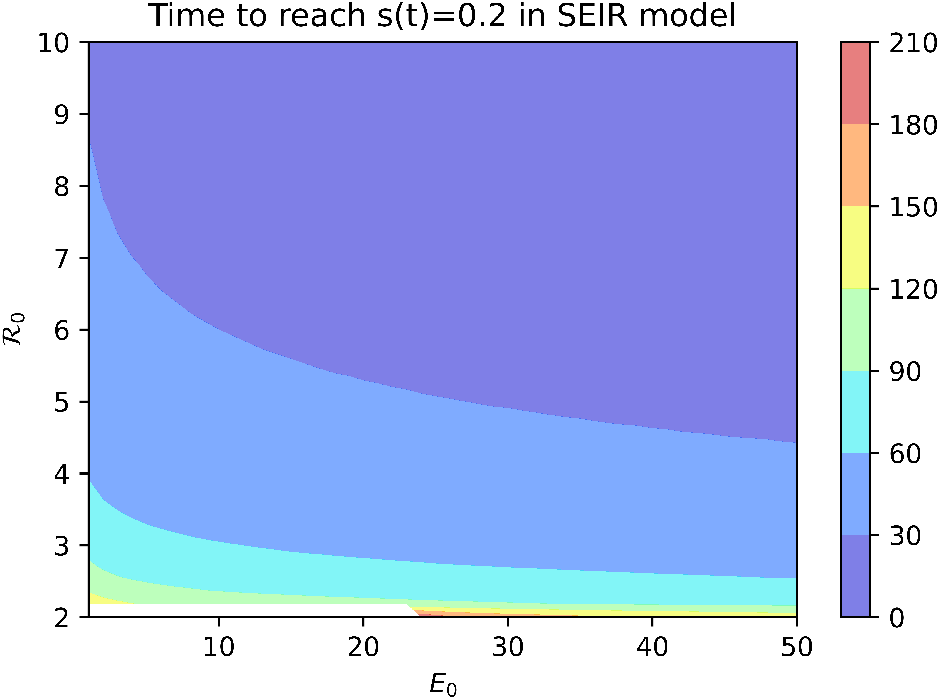
Time to infect 80% of population in SEIR model with median incubation period of 5.1 days [12] and mean infectious period of 5.6 days.

An alternative explanation is that the outbreak involved more than one initially infected prisoner. From Figure 2, we see that for a fixed *ℛ*_0_ value, increasing *E*_0_ decreases the time needed to infect 80% of the population. However, an initial condition of *E*_0_ *>* 471 is needed for an outbreak with *ℛ*_0_ = 3.27 to infect 80% of the population within three weeks.

### Time of initial outbreak circulation

A third possibility is that SARS-CoV-2 was circulating among prisoners prior to April 3. Figure 3 shows the log-likelihood (2.6) for observing the mass testing results in MCI according to the SEIR model (2.1) as a function of *ℛ*_0_ and the outbreak onset date, with *E*_0_ fixed at one. The outbreak onset date and *ℛ*_0_ are indistinguishable from the mass testing data alone, with the yellow “wishbone” in Figure 3 corresponding to outbreak onset and *ℛ*_0_ pairs that are equally likely given the data. Outbreak onsets in late March or later correspond to *ℛ*_0_ values larger than ten, while earlier outbreak onsets correspond to smaller *ℛ*_0_ values. Note that onset dates prior to March are required to give *ℛ*_0_ values of less than five.

**Figure 3:**
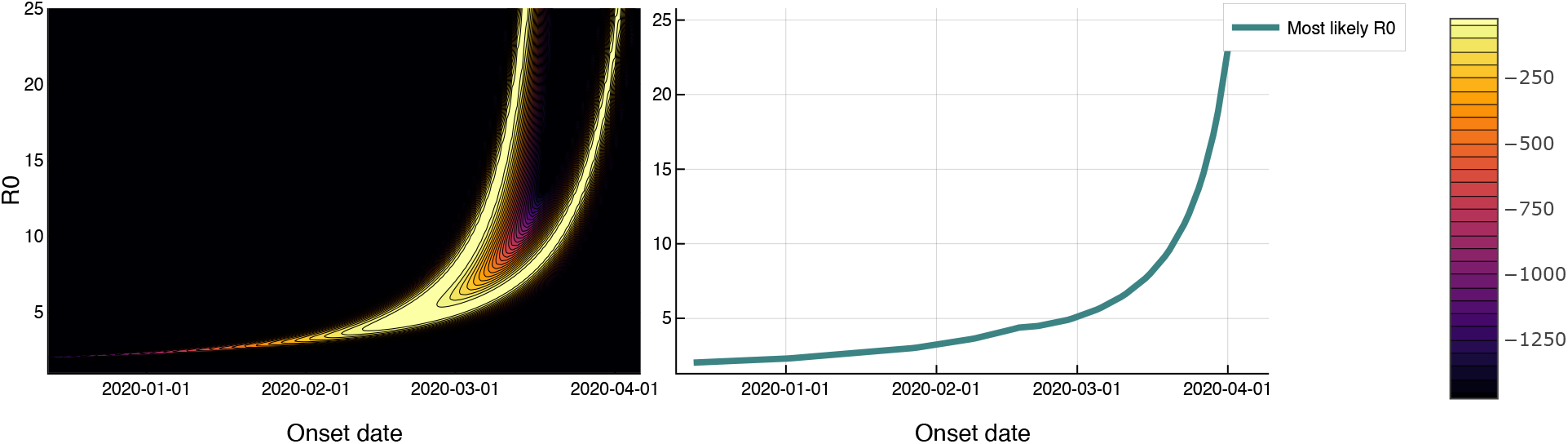
*Left*. Log-likelihood in the *ℛ*_0_-outbreak onset date plane, for mass testing data under a SEIR model with three week test-positive window following infectious onset. *Right. ℛ*_0_ values that maximize the likelihood as a function of outbreak onset date. This curve corresponds to following the right-most branch of the ‘wishbone’ in the likelihood plot.

## 4 Discussion

The official reports from ODRC describe widespread infection of MCI inmates with SARS-CoV-2 within the span of three weeks. Our analysis indicates three (non-exclusive) possible explanations for this rapid spread: (i) values for the basic reproduction number that are far higher than the *ℛ*_0_ values between two and three that have been estimated for non-prison settings in the United States [13], (ii) initial exposure of a large number of infected prisoners, consistent with an extreme super-spreading event, and (iii) early, undetected circulation of SARS-CoV-2 among prisoners prior to April 3. All three possibilities speak to the vulnerabilities of the prison population for COVID-19. The permissive conditions for spread within correctional facilities, the challenges for disease surveillance and care in these settings, and the inextricable link between COVID-19 within correctional facilities and disease spread in the broader community, have been eloquently discussed by others [4–8]. None of these issues can be adequately addressed without collection and analysis of data regarding COVID-19 in prisons. Officially reported data such as those considered here can already be used for statistical and mathematical analyses [14, 15]. Further data collection and dissemination, for example distinguishing symptomatic versus mass testing and including both positive and negative test results, would facilitate additional analyses that would deepen our understanding of transmission. We urge health departments and corrections departments to undertake collection of accurate data and to make these data available for analysis. These efforts are imperative for preventing COVID-19 spread in prisons.

## Data Availability

Code for this project is available at https://github.com/wasiur/PrisonCOVID19Analysis.

## Footnotes

1 The July 21 ODRC report [2] reports that 2023 inmates had recovered from COVID-19. We take the May 5, 2020 listing of 2453 inmates at MCI as the denominator.

2 As stated in the publicly available ODRC report from March 30, 2020: “Based on a staff member reporting a positive COVID test, MCI is operating under modified movement and the population is being separated by unit along with other precautionary measures. Every inmate at MCI is monitored daily and has their temperature taken along with a check for symptoms. Currently, there are no inmates symptomatic for COVID-19.”

3 These numbers come directly from data on the ODRC website. There is a lag of a few days between when mass testing occurred (April 16) and when jumps in case counts are reported in the ODRC data (April 18 and April 19 for inmates; April 20 for staff). This lag may reflect delay in data entry.

